# Quality Assurance Assessment of Intra-Acquisition Diffusion-Weighted and T2-Weighted Magnetic Resonance Imaging Registration and Contour Propagation for Head and Neck Cancer Radiotherapy

**DOI:** 10.1101/2021.12.13.21267735

**Authors:** Mohamed A. Naser, Kareem A. Wahid, Sara Ahmed, Vivian Salama, Cem Dede, Benjamin W. Edwards, Ruitao Lin, Brigid McDonald, Travis C. Salzillo, Renjie He, Yao Ding, Moamen Abobakr Abdelaal, Daniel Thill, Nicolette O’Connell, Virgil Willcut, John P. Christodouleas, Stephen Y Lai, Clifton D. Fuller, Abdallah S. R. Mohamed

## Abstract

**Background/Purpose:** Adequate image registration of anatomic and functional MRI scans is necessary for MR-guided head and neck cancer (HNC) adaptive radiotherapy planning. Despite the quantitative capabilities of diffusion-weighted imaging (DWI) MRI for treatment plan adaptation, geometric distortion remains a considerable limitation. Therefore, we systematically investigated various deformable image registration (DIR) methods to co-register DWI and T2-weighted (T2W) images.

**Materials/Methods:** We compared three commercial (ADMIRE, Velocity, Raystation) and three open-source (Elastix with default settings [Elastix Default], Elastix with parameter set 23 [Elastix 23], Demons) post-acquisition DIR methods applied to T2W and DWI MRI images acquired during the same imaging session in twenty immobilized HNC patients. In addition, we used the non-registered images (None) as a control comparator. Ground truth segmentations of radiotherapy structures (tumor and organs at risk) were generated by a physician expert on both image sequences. For each registration approach, structures were propagated from T2W to DWI images. These propagated structures were then compared with ground truth DWI structures using the Dice similarity coefficient and mean surface distance.

**Results:** 19 left submandibular glands, 18 right submandibular glands, 20 left parotid glands, 20 right parotid glands, 20 spinal cords, and 12 tumors were delineated. Most DIR methods took < 30 seconds to execute per case, with the exception of Elastix 23 which took ∼458 seconds to execute per case. ADMIRE and Elastix 23 demonstrated improved performance over None for all metrics and structures (Bonferroni-corrected p < 0.05), while the other methods did not. Moreover, ADMIRE and Elastix 23 significantly improved performance in individual and pooled analysis compared to all other methods.

**Conclusions:** The ADMIRE DIR method offers improved geometric performance with reasonable execution time so should be favored for registering T2W and DWI images acquired during the same scan session in HNC patients. These results are important to ensure the appropriate selection of registration strategies for MR-guided radiotherapy.

## 1. Introduction

Radiation therapy (RT) is an essential treatment modality for head and neck cancer (HNC) ^1^. Conventionally, RT has relied on radiographic images to enable pre-treatment segmentation of target volumes and nearby organs at risk (OAR) to plan intensity-modulated doses ^2, 3^. However, throughout RT, the dynamic changes in target volumes and OARs and patient-specific changes (e.g., weight loss) can lead to unintended doses of radiation to OARs and subsequent debilitating side effects ^4^. These potential unintended doses are particularly relevant for HNC because the head and neck region is home to various complex, highly radiosensitive structures and tissue interfaces that can drastically change during RT ^4, 5^.

Image-guided RT, during which radiation dose can be administered in tandem with onboard imaging, has become a promising alternative to intensity-modulated RT, in part due to increasingly ubiquitous image-guided technology, such as MR-Linac devices ^6, 7^. MR-guided treatment also affords the ability to capture distinct patient anatomy with varying contrasts via weighted sequence acquisitions, such as T2-weighted (T2W) images, and functional information, such as through diffusion-weighted imaging (DWI). DWI has shown particular benefit in aiding treatment adaptation through improved detection of target volumes and assessment of treatment response ^8^. Therefore, combined T2W and DWI acquisition enable the gathering of anatomic and functional information that can be used for adaptive MR-guided personalized RT.

Anatomical and functional sequences acquired in the same imaging session for MR-guided treatment often have minimal variation in patient position and geometry between sequence acquisitions, often due to careful patient immobilization ^9^. However, these multisequence acquisitions can be misaligned by motion artifacts from respiration or swallowing ^4^, susceptibility artifacts, chemical shift artifacts, ghosting artifacts ^8^, and geometric distortions ^10^. Post-acquisition image registration, the process by which homologous image voxels from multi-temporal or multi-modal image sets are mapped to each other ^11, 12^, is an important approach to align anatomical and functional sequences. Rigid image registration involves global matching between image sets, while deformable image registration (DIR) uses optimization algorithms to adjust image transformation models. Most implementations of DIR involve a transformation that establishes a geometric correspondence between fixed and moving images, an objective function, and an optimization approach to maximize the similarity between images ^13–15^. Importantly, even minor differences in patient anatomy can result in devastating dose administration in HNC ^4, 16^, highlighting the need for consistent image co-registration when propagating segmentations of target volumes and OARs for radiotherapy treatment planning. Therefore, determining the impact of post-acquisition registration techniques (i.e., DIR) on multisequence MRI acquisitions is crucial for MR-guided treatment of HNC.

While we have previously investigated intra-modality CT to CT registration ^17^ and inter-modality CT to MRI registration ^18^, to our knowledge, there are no studies that investigate registration techniques for intra-acquisition MRI in HNC. Therefore, to facilitate further development and optimization of MR-guided RT adaptive planning technologies, we systematically analyzed DIR methods in T2W and DWI MRI sequences acquired during the same imaging session.

## 2. Methods

We developed a quality assurance workflow for evaluating and benchmarking the performance of different image registration methods for T2W and DWI images (described in subsection 2.2) of 20 HNC patients (2.1). We evaluated different DIR methods provided by the commercial RT treatment planning software, as well as open-source DIR implementations (2.3). The deformation vector field (DVF) generated by each method was used to propagate the manually segmented structures (2.2) from T2W to DWI images. The structures propagated by different methods to DWI images were compared to the ground truth segmentations for performance evaluation (2.4).

### 2.1. Patient Characteristics

Twenty patients with HNC who had undergone RT in a clinical trial (NCT03145077) were included in this analysis. All clinical and imaging data were generated between May 30, 2017 and April 1, 2019 and were retrospectively collected under a HIPAA-compliant protocol (PA16-0302) that was approved by The University of Texas MD Anderson Cancer Center’s institutional review board. All patients provided study-specific informed consent. The median patient age was 54 years, with a male predominance (80%). Primary tumor sites included the oropharynx, nasopharynx, and oral cavity. Full patient clinical and demographic characteristics are summarized in **Table 1**.

**Table 1.**
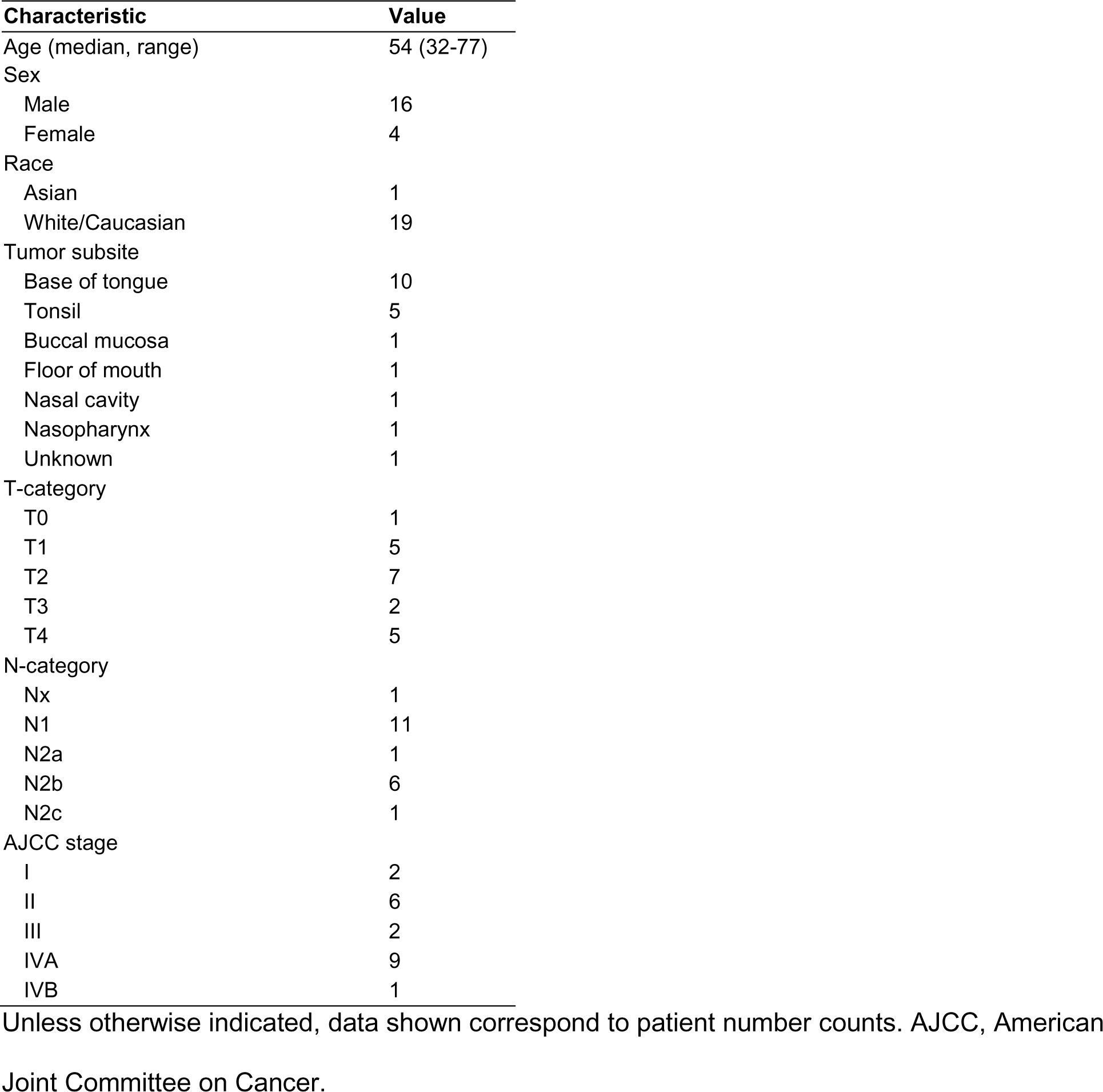
Patient clinical and demographic characteristics.

### 2.2. Imaging Data

Pre-RT T2W and DWI MRI sequences in Digital Imaging and Communications in Medicine (DICOM) format for each of the 20 patients were curated from our imaging databases. T2W images and DWI images with a b value of 0 were collected in the same imaging session while the patient was immobilized in a thermoplastic mask using a 1.5

Tesla Siemens MRI simulator. Characteristics of the imaging sequences are shown in **Table 2**. For each image set (T2W image and DWI image), ground truth segmentations for the left and right submandibular glands, left and right parotid glands, cervical spinal cord, and primary gross tumor volume were manually generated by a trained physician expert (radiologist with > 5 years of experience in HNC). All segmentations were generated in Velocity AI (v.3.0.1; Varian Medical Systems; Palo Alto, CA, USA) in DICOM RT structure format. The anonymized image sets and structure files are publicly available online through Figshare (10.6084/m9.figshare.17162435, under embargo until manuscript acceptance).

**Table 2.** MRI sequence acquisition parameters. Median value displayed with range of values shown in parenthesis. No parenthesis indicates all values were the same for all patients.

| Acquisition Parameter | T2W | DWI |
| --- | --- | --- |
| Repetition time (ms) | 4800 | 5000 (1500-7000) |
| Echo time (ms) | 80 | 65 (50-102) |
| Echo train length | 15 | 15 (0-63) |
| Flip angle (°) | 180 (166-180) | 120 (90-180) |
| Slice thickness (mm) | 2 | 4 |
| In-plane resolution (mm) | 0.5 | 2.0 (1.0-2.0) |
| Slice gap (mm) | 2 | 4 |
| Acquisition matrix | 256x230 | 128x128 |
| Pixel bandwidth (Hz/px) | 300 | 870 (750-1220) |
| Number of averages | 1 | 2 |
| Number of axial slices | 120 | 28 (24-48) |

T2W, T2-weighted magnetic resonance imaging; DWI, diffusion-weighted magnetic resonance imaging.

### 2.3. Image Registration

For this analysis, we investigated several DIR registration methods from different commercial radiotherapy software packages and open-source implementations. Specifically, the following DIR methods were utilized:

1. ADMIRE: A proprietary approach from the commercial software package ADMIRE (v.3.29; Elekta AB; Stockholm, Sweden) that implements an atlas-based approach with head pose correction, dense mutual-information, and refinement using a deformable surface model.
2. Velocity: A proprietary approach from the commercial software package Velocity AI (v.3.0.1; Varian Medical Systems; Palo Alto, CA, USA) that implements a 3-pass (coarse-medium-fine resolution) modified B-spline.
3. Elastix Default: An open-source approach from the popular medical image registration library Elastix (SimpleElastix Python interface ^19^) that utilizes a multi-resolution B-spline (default parameter map). Specifically, the algorithm uses an adaptive stochastic gradient descent optimizer, linear interpolator, FixedImagePyramidSchedule = 8 8 8 4 4 4 2 2 2 1 1 1, MovingImagePyramidSchedule = 8 8 8 4 4 4 2 2 2 1 1 1, number of resolutions = 4, and 4096 spatial samples. More information on the algorithm can be found in the Elastix documentation ^20^.
4. Elastix 23: An open-source approach from the popular medical image registration library Elastix (SimpleElastix Python interface ^19^) that utilizes localized mutual information combined with a bending energy penalty in a B-spline transformation (referred to as parameter map 23 in the Elastix Zoo) ^21^. Specifically, the algorithm uses an adaptive stochastic gradient descent optimizer, B-spline interpolator, Image Pyramid Schedule = 8 8 2 4 4 1 1 1 0.5, number of resolutions = 3, and 10000 spatial samples. This algorithm was selected because it was explicitly developed for multi-modality head and neck registration. More information on the algorithm can be found in the Elastix documentation ^20^.
5. Demons: An open-source approach based on the Demons ^22^ family of algorithms available in SimpleITK ^23^. Specifically, the algorithm uses a multi-resolution framework with shrink factors of [4,2,1] and smoothing sigmas of [8,4,0], a linear interpolator, and a gradient descent optimizer (learning rate = 1.0, number of iterations =20, convergence minimum value = 1e^-6^, convergence window size = 10).
6. Raystation: A proprietary approach from the commercial software package RayStation Research (RaySearch Laboratories, Stockholm, Sweden) that implements a modified ANAtomically CONstrained Deformation Algorithm ^24^ using only intensity-based registration.

For all cases, the DWI image was used as the fixed image, and the T2W image was used as the moving image; T2W images were resampled to the DWI image. To maintain adequate comparisons between structures generated on T2W images and DWI images, before registration all structures were cropped to the image with the smaller field of view, e.g., DWI image. For additional deformable vector field (DVF) visualization and analysis, Jacobian determinant matrices were derived from DVF files of each method using SimpleITK or as direct output from ADMIRE. As a control comparator for all cases, we also analyzed the raw images with no post-acquisition registration applied, i.e., an implicit rigid registration as a byproduct of patient immobilization (labeled as “None”). After the registration process, for each method, we propagated the ground truth segmentations from the T2W images to the DWI images using the corresponding transformations to generate propagated structures (**Figure 1A-H**). These propagated structures were then compared to the ground truth structures on the DWI image in the subsequent analysis. Before the analysis, all images and structure files were transformed into Neuroimaging Informatics Technology Initiative format. Finally, because there were small variations in the inferior and superior slices of the cervical spinal cord, we cropped these structures so that the heights of the propagated segmentation and ground truth segmentation were equal (**Figure 1I)**.

**Figure 1.**
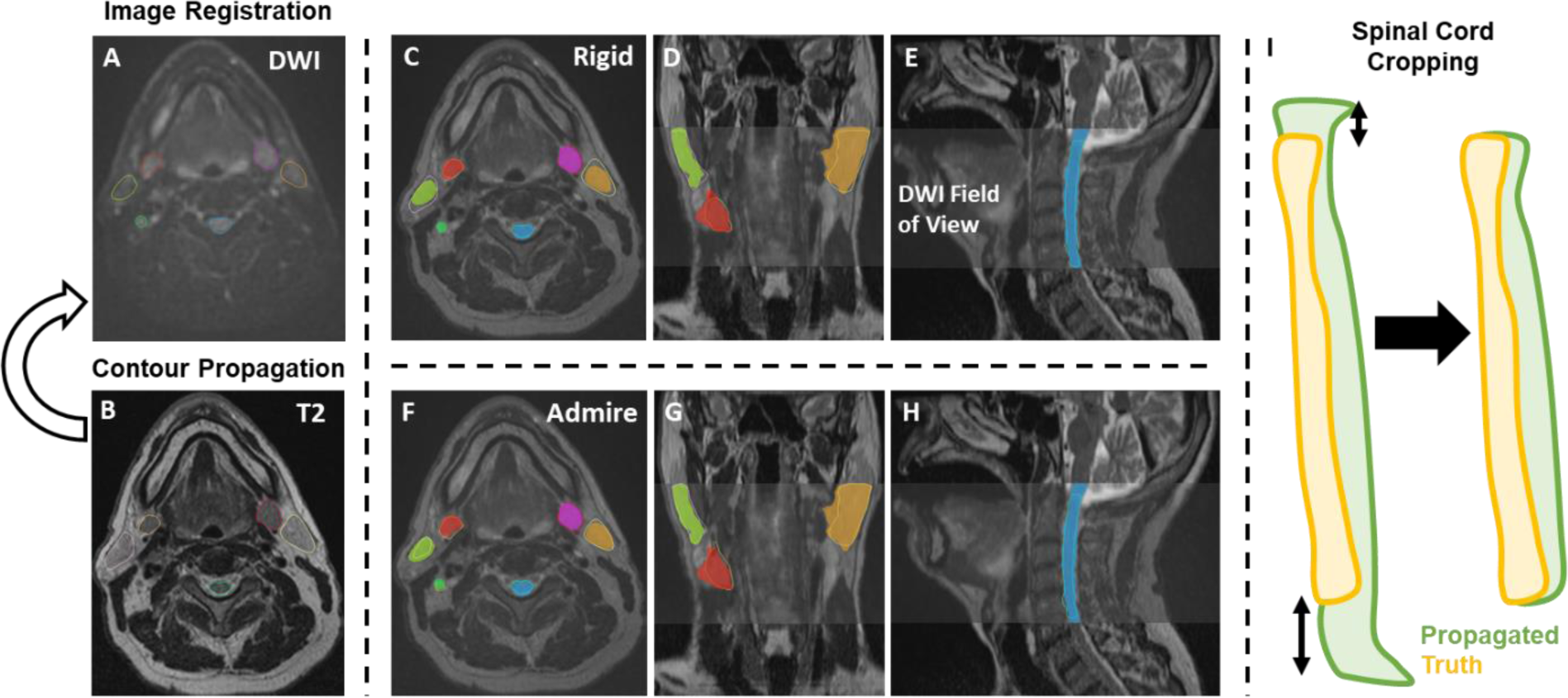
Study workflow. Contours are propagated from the moving image (**B**, T2-weighted image [T2]) to the fixed image (**A**, diffusion-weighted image [DWI]) for each registration method. **C-E** and **F-H** show propagated and ground truth structures for the implicit rigid and ADMIRE approaches, respectively. The spinal cord was also cropped so that the height of the propagated segmentation and ground truth segmentation were equal (**I**).

### 2.4. Statistical Analysis

Several evaluation metrics were used to compare the propagated structure sets after registration to the ground truth structures delineated on the DWI images. Specifically, for each individual structure, the Dice similarity coefficient (DSC) and mean surface distance (MSD) were calculated as they are well-established and ubiquitous metrics for measuring volumetric and surface distance information, respectively ^25^. Additional volumetric and surface distance metrics were calculated for supplementary analyses (**Appendix A**). Metrics were calculated using the surface-distance Python package ^26^ and in-house Python code. After performing a Shapiro-Wilk test ^27^, we found that our data were not normally distributed (p<0.05). Therefore, we used nonparametric statistical tests for our analysis. For each metric and each structure, we compared registration methods against ‘None’ using one-sided Wilcoxon signed-rank tests (alternative hypothesis of greater than the null hypothesis for DSC and alternative hypothesis of less than the null hypothesis for MSD) with Bonferroni adjustments for multiple comparisons ^28^. Similarly, we pooled metrics for OARs for sub-analysis and performed pair-wise analysis using previously described Wilcoxon signed-rank tests with Bonferroni corrections. Interobserver variability, dosimetric, target registration error (TRE) for a subset of 5 patients, and DVF Jacobian matrix analysis were performed in **Appendix B**, **Appendix C**, **Appendix D,** and **Appendix E**, respectively. For all statistical analyses, p-values less than 0.05 were considered significant. All statistical analyses were performed in Python v.3.7 ^29^.

## 3. Results

### 3.1. Quantitative Comparison

19 left submandibular glands, 18 right submandibular glands, 20 left parotid glands, 20 right parotid glands, 20 cervical spinal cords, and 12 primary tumors in both T2W and DWI images were used in the analysis. ADMIRE, Velocity, Elastix Default, Elastix 23, Demons, and Raystation each took approximately 23, 7, 46, 458, 4, and 13 seconds to complete one case, respectively. For each method, most OAR structures had similar performance across the multiple metrics except for the cervical spinal cord which was notably worse (**Figure 2**); we therefore performed additional analysis to investigate spinal cord structures in individual cases in **Appendix F**. Compared to the structures generated by no registration (“None”), all structures demonstrated an improvement with the ADMIRE and Elastix 23 methods, and worsened with all other methods (**Figure 2**). Specifically, the ADMIRE and Elastix 23 methods showed significant improvements (p<0.05 on one sided Wilcoxon signed rank test) for both DSC and MSD metrics for all structures (**Figure 3**). When metrics were pooled across structures, similar trends emerged where the ADMIRE and Elastix 23 methods demonstrated the best performance compared to the other methods, with DSC gains over None of up to .05 and .07, respectively in the OARs, and up to .08 and .09, respectively, in the tumor (**Table 3**). Moreover, pair-wise comparisons of pooled OAR structures and the tumor demonstrated that Elastix 23 offered significantly improved performance over ADMIRE (p<0.05), and that Elastix Default, Demons, and Raystation were consistently outperformed by the other methods (**Figure 4**). Dosimetric (**Appendix C**) and TRE (**Appendix D**) analysis revealed no significant improvements of any DIR methods compared to None.

**Figure 2.**
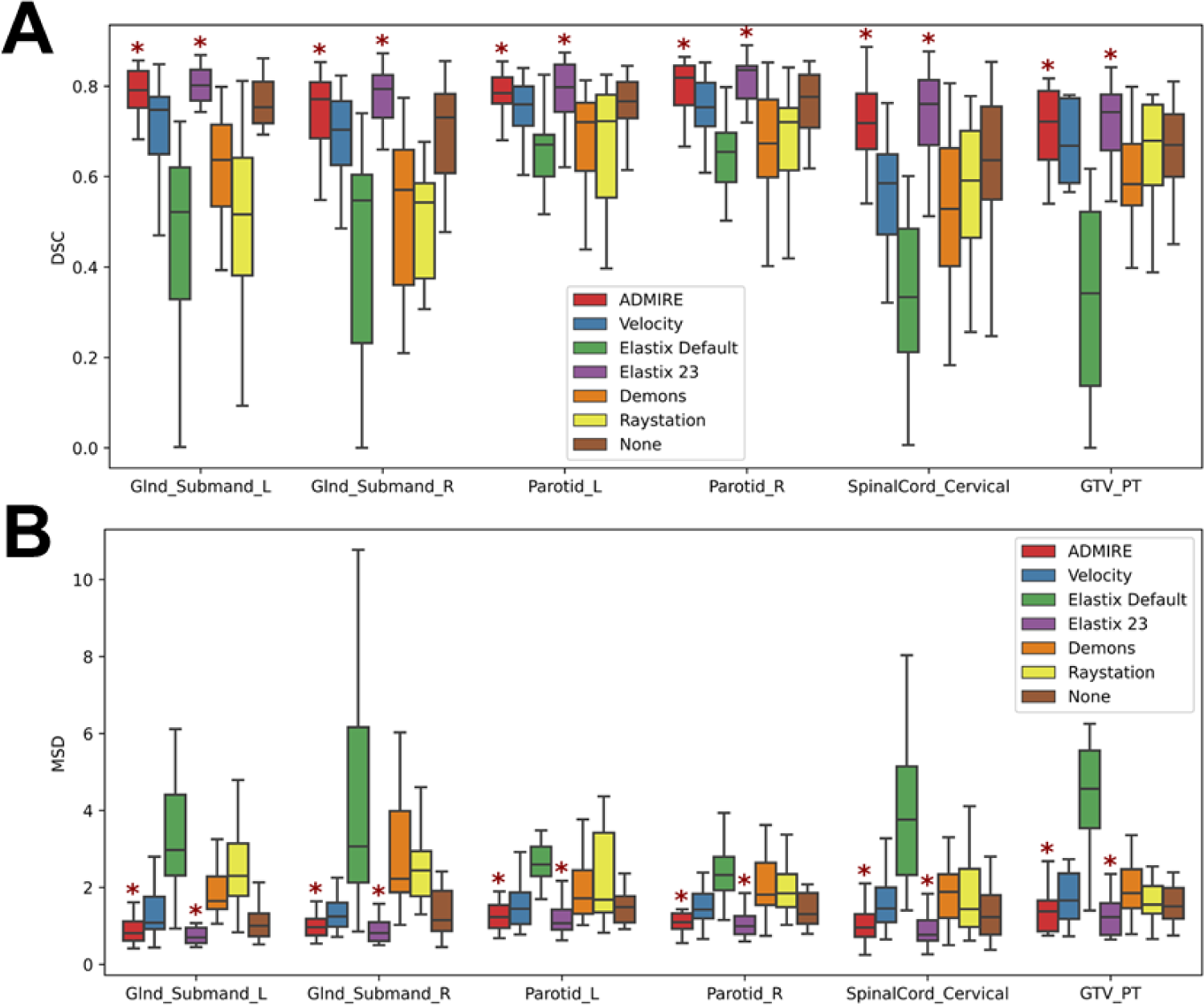
Box plots of evaluation metrics for each structure according to the registration method for Dice similarity coefficient (DSC) [**A**] and mean surface distance (MSD) [**B**]. Asterisks indicate a significant improvement between the registration method and no registration (None). Glnd_Submand, submandibular gland; L, left; R, right; GTV_PT, primary gross tumor volume.

**Figure 3.**
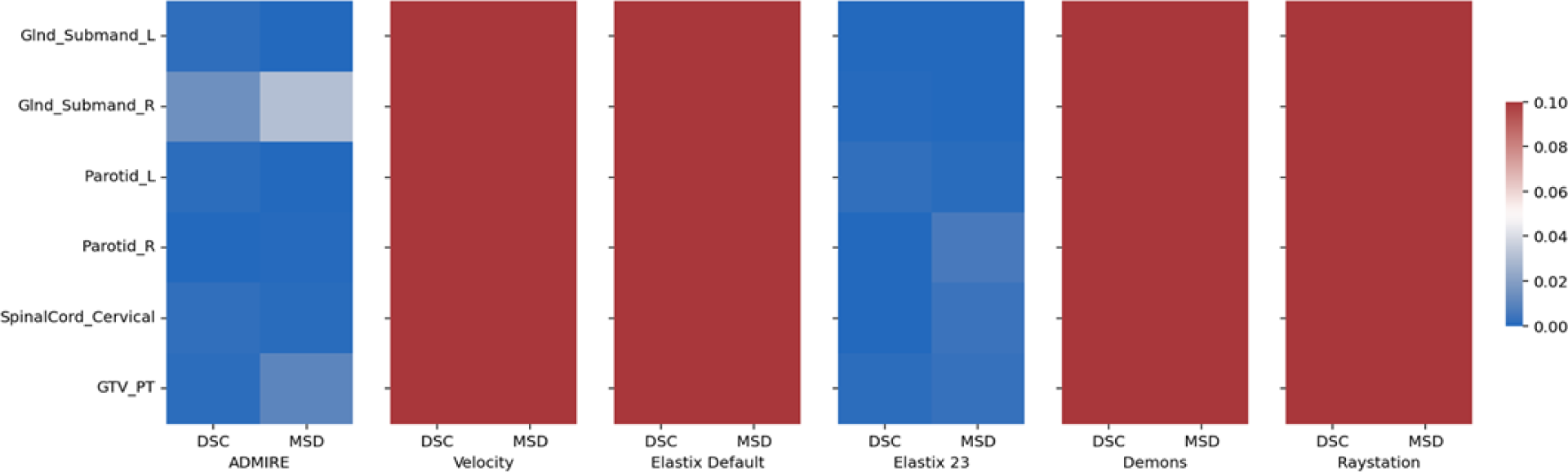
Heatmap of Bonferroni corrected p-values for one-way Wilcoxon-signed rank tests between various registration methods and no registration (None) indicating significant improvement across evaluation metrics and structures. Blue colors correspond to significant p-values (p<0.05) while red colors correspond to non-significant values (p>0.05). Glnd_Submand, submandibular gland; L, left; R, right; GTV_PT, primary gross tumor volume; DSC, Dice similarity coefficient; MSD, mean surface distance.

**Figure 4.**
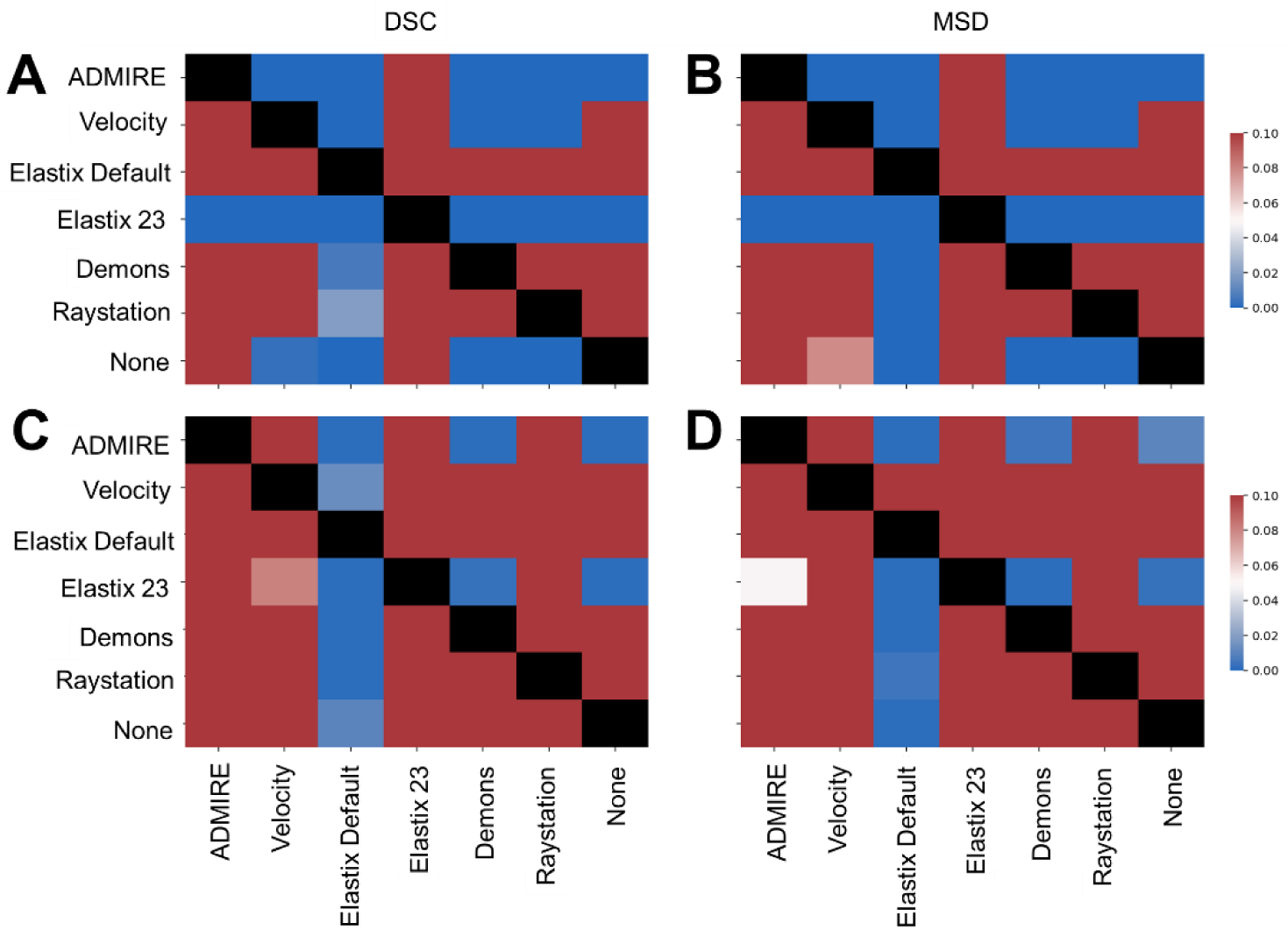
Heatmap of Bonferroni corrected p-values for one-way Wilcoxon-signed rank tests for pair-wise comparisons between various registration methods indicating significant improvement of method in row vs. method in column. Top plots correspond to all pooled organ at risk structures using Dice Similarity Coefficient (DSC) (**A**) and mean surface distance (MSD) (**B**). Bottom plots correspond to tumor using DSC (**C**) and MSD (**D**). Blue colors correspond to significant p-values (p<0.05) while red colors correspond to non-significant values (p>0.05). Comparisons between the same method (diagonal entries) are blacked out.

**Table 3.** Evaluation metrics (mean ± standard deviation) across pooled structures according to each registration method. DSC, Dice similarity coefficient; MSD, mean surface distance.

|  | Metric | ADMIRE | Velocity | Elastix Default | Elastix 23 | Demons | Raystation | None |
| --- | --- | --- | --- | --- | --- | --- | --- | --- |
| OARs | DSC | 0.76±0.09 | 0.67±0.16 | 0.51±0.21 | 0.78±0.08 | 0.60±0.16 | 0.58±0.18 | 0.71±0.13 |
| | MSD (mm) | 1.12±0.51 | 1.68±1.25 | 3.68±2.92 | 1.01±0.49 | 2.19±1.16 | 2.39±1.92 | 1.46±0.92 |
| Tumor | DSC | 0.70±0.09 | 0.59±0.25 | 0.33±0.20 | 0.71±0.09 | 0.60±0.12 | 0.63±0.16 | 0.62±0.21 |
| | MSD (mm) | 1.38±0.55 | 3.63±6.62 | 5.49±3.59 | 1.25±0.52 | 2.00±0.82 | 1.88±1.03 | 2.64±3.56 |

OARs, organs at risk; DSC, Dice similarity coefficient; MSD, mean surface distance.

### 3.2. Qualitative Comparison

We visually compared the deformed T2W images of the various DIR methods and their corresponding Jacobian determinant matrices in **Figure 5**. Generally, most methods often yielded visually similar deformed image outputs as the ground-truth T2W image. However, large deviations in DVF warping could sometimes be observed for Velocity and Elastix Default. Additional quantitative analysis of Jacobian determinants for the various methods can be found in **Appendix E**.

**Figure 5.**
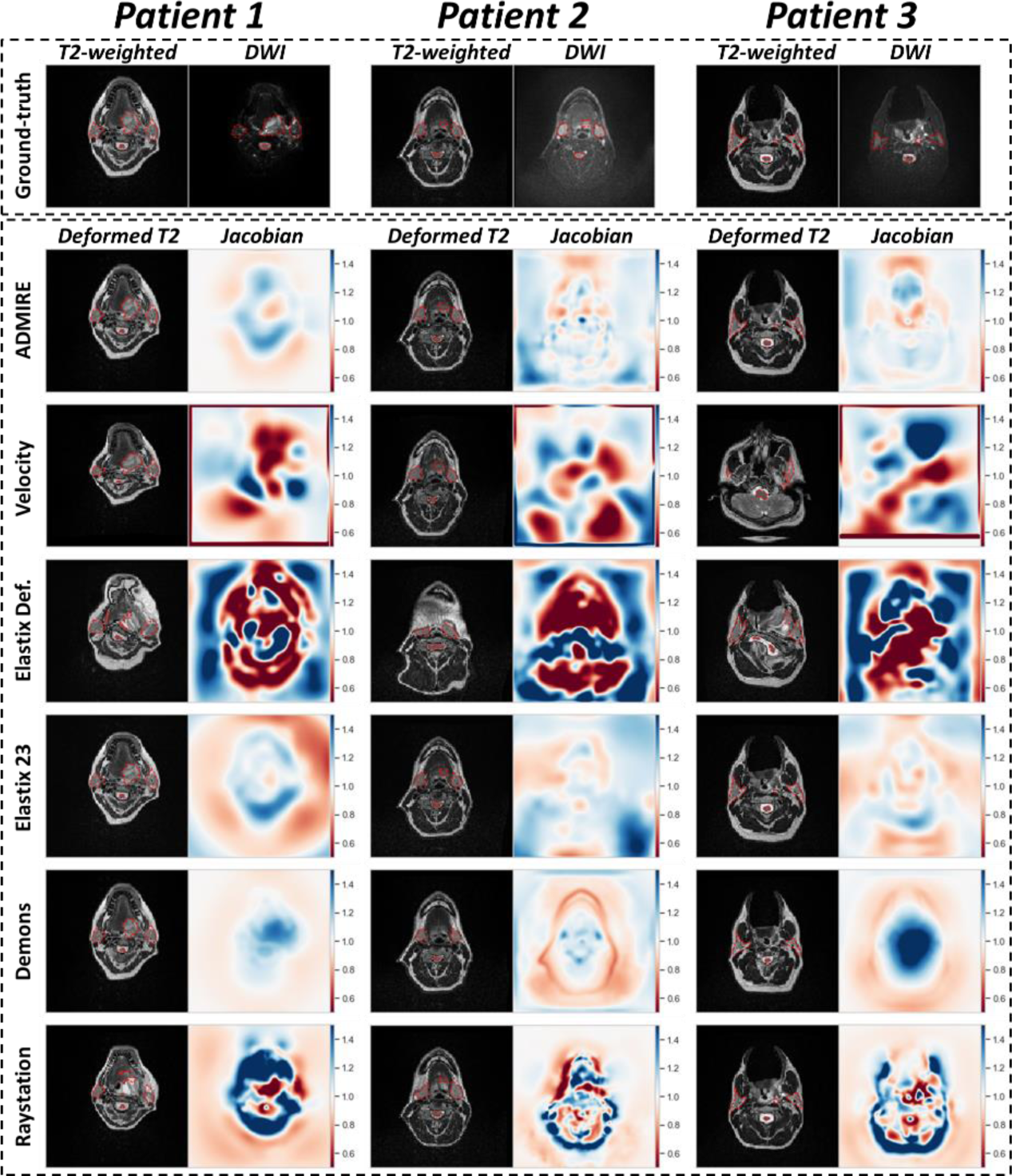
Visualization of deformable image registration method outputs for 3 example patients. The top row shows original T2-weighted image and diffusion weighted image (DWI) with ground-truth segmentations overlaid (red dotted outline). The deformed T2-weighted image with overlaid propagated segmentation (red dotted outline) and corresponding Jacobian determinant is shown for each image registration method. Jacobian determinants greater than 1 indicate volume expansion, between 0 and 1 indicate volume reduction, and equal to 1 indicates no change.

## 4. Discussion

In this study, we systematically analyzed a variety of DIR methods and compared them to non-registered images from multisequence MRI acquisitions for HNC image-guided treatment applications. Our results highlight that specific DIR methods can improve upon pre-established head and neck immobilization, as shown by measuring the similarity of propagated ground truth segmentations from T2W images to DWI images compared to ground truth segmentations on DWI images (**Figures 2 and 3**).

The best overall results were obtained using ADMIRE and Elastix 23 (**Table 3**), with all metric and structure combinations having significantly better performance than None (**Figure 3**). While we tested other deformable methods (i.e., Velocity, Demons, Elastix Default, and Raystation), they were significantly worse for most metric and structure combinations when compared to the None. Moreover, Velocity and Elastix Default sometimes yielded implausible DVFs, as indicated by qualitative and quantitative analysis of Jacobian determinants (**Figure 4, Appendix E**), which may be due to these DIR algorithmic implementations being unable to accommodate large variations in intensity domains of the T2W and DWI images. Importantly, almost all structures individually and on pooled analysis showed improved DSC and MSD for the ADMIRE and Elastix 23 methods (**Table 3**). These results indicate that these methods provide significantly improved volumetric and surface distance overlap, which may warrant their use during intra-acquisition MRI sequences for MR-guided treatment. Moreover, while dosimetrically there were no significant improvements for any structures for ADMIRE and Elastix 23 compared to None (**Appendix C**), these differences may still be clinically significant. Similarly, for TRE analysis on a subset of methods, ADMIRE often offered decreased registration error compared to no registration (**Appendix D**), but was nonsignificant, likely secondary to the already minimal registration errors induced through the use of an immobilization mask. Notably, Elastix 23 provided significantly improved volumetric and surface distance performance compared to ADMIRE. However, these improvements may not be clinically significant (DSC gains of ∼1%) and come at the cost of a much longer execution time (∼7 minutes longer), therefore, ADMIRE should likely be preferred for workflows where time is a limiting factor, i.e., adaptive radiotherapy. It is also worth noting the spinal cord is especially sensitive to distortion-causing artifacts ^30^, making it a particularly challenging structure to co-register adequately. While the general performance for the spinal cord was lower than that of other OAR structures, the ADMIRE based methods were still able to offer significantly improved performance compared to the implicit rigid registration (**Figures 2 and 3)**; cases with lower performance tended to have a larger degree of spinal curvature than cases with higher performance (**Appendix F**). Therefore, while the ADMIRE method should still be preferred over no registration, special caution should be used in quality assurance when used for spinal cord segmentations. Notably, all estimated metrics between any registration method and None showed no significant differences using segmentations generated by different observers (**Appendix B**), indicating that our data are not confounded by interobserver variability.

While several previous studies have investigated the relative performance of registration methods in various anatomical sites ^31–33^, there is a general lack of investigations of head and neck imaging. However, a few recent important studies have investigated registration quality assessment in head and neck imaging using radiotherapy structure analysis similar to our current study ^17, 18^. For example, Mohamed et al. ^17^ investigated the registration quality of diagnostic CT to simulation CT in HNC where images were acquired at different time points and with different scan settings and found that certain DIR methods demonstrated improved performance over a control group (rigid registration) for OAR and target conformance for most comparison metrics, similar to our study. Oppositely, Kiser et al. ^18^ showed that for CT and T2W MRI scans acquired with standard treatment immobilization techniques, MRI to CT DIR was not superior to rigid registration, with neither technique producing clinically satisfactory results (DSCs of 0.62 - 0.65). Importantly, the ADMIRE method investigated in our study produce potentially clinically meaningful results as we observe significant performance gains across various structures that may impact MR-guided treatments with a reasonable execution time.

To date, no systematic anatomical to functional MRI registration studies have been performed for HNC. However, intra-acquisition MRI registration techniques have been investigated in other anatomical sites. Specifically, several studies have compared registration techniques for various MRI sequences in the prostate ^34–36^. For example, Buerger et al. compared the performance of five state-of-the-art DIR image registration techniques for accurate image fusion of DWI with T2W images and found fast elastic image registration provided improved performance compared to other deformable techniques such as B-spline and Demons ^35^. This result was further echoed in Eriksson et al., which confirmed that fast elastic image registration was the best technique for T1-weighted to T2W anatomic sequence registration ^36^. Our results are consistent with these observations that selecting appropriate deformable techniques offers significantly improved performance for intra-acquisition registration.

There are several limitations to our study. We limited our analysis of intra-acquisition registration techniques in MRI to T2W and DWI sequences since these are the most germane to current MR-guided RT applications. However, several additional sequences can be studied to investigate these phenomena. In this study, we only tested b0 images, which were readily available and common for DWI workflows in HNC. Moreover, as no images suffered from major geometric distortion, we did not address geometric distortion in this study. Future iterations of this study should investigate other DWI-derived images and the influence of geometric distortion on DIR. Notably, we have found dosimetric improvements for certain DIR methods (though non-significant), but the clinical significance of these improvements is unknown and should be confirmed through additional experiments such as normal tissue complication probability calculations. Additionally, we have limited our investigation to a few critical RT HNC structures of interest using one expert observer; future studies should investigate a greater number of structures in a greater number of patients with a larger number of observers. It should also be noted that while most contoured structures were not expected to vary considerably between T2 and DWI, pathologic structures may be interpreted differently on these images, thus caution should be used when interpreting results for the tumor. Finally, we have limited our analysis to intra-acquisition images collected during the same image acquisition session. However, for MR-guided RT applications, registration techniques are also relevant for images taken at different time points. Therefore, future studies should investigate these registration techniques applied to different imaging time points in an MR-guided RT workflow.

## 5. Conclusions

In summary, this is the first study to investigate intra-acquisition MRI registration quality in HNC patients. We identify a deformable registration technique from the ADMIRE software package that offers the most significant gains in registration quality with reasonable execution time for T2W to DWI image registration compared to other methods. Our results are a crucial first step towards registration quality assurance for MR-guided treatment approaches that implement multi-sequence acquisitions combining anatomical and functional imaging.

## Data Availability

The anonymized image sets and structure files are publicly available online through Figshare (10.6084/m9.figshare.17162435). Data under embargo until acceptance in peer-reviewed journal.

## Acknowledgments

The authors thank Ann Sutton, Scientific Editor, and Ashli Nguyen-Villarreal, Associate Scientific Editor, in the Research Medical Library at The University of Texas MD Anderson Cancer Center, for editing this article. The authors also acknowledge the following people for their contributions to the NIH-funded academic-industrial partnership grant (R01DE028290) that funded this work and for their general support and feedback regarding this project: Spencer Marshall, Hafid Akhiat, Michel Moreau, Nathan Cho, Edyta Bubula-Rehm, Chunhua Men, and Etienne Lessard of Elekta and Alex Dresner of Philips.

## Abbreviations

T2W: T2-weighted
DWI: diffusion-weighted imaging
RT: Radiation therapy
HNC: head and neck cancer
OAR: organs at risk
DIR: deformable image registration

## Appendices

## Appendix A: Additional Metric Evaluations

For each individual structure, in addition to the Dice similarity coefficient (DSC) and the mean surface distance (MSD) as reported in the main text, we also calculated the following evaluation metrics: false-negative DSC (FN-DSC), false-positive DSC (FP-DSC), surface DSC (S-DSC), 95% Hausdorff distance (95% HD), and mean surface distance (MSD). For S-DSC, a tolerance of 2.5 mm was selected as a suitable tolerance from previous studies ^1, 2^. Boxplot representations are shown in **Figure A1**, while significance test heatmaps are shown in **Figure A2**.

**Figure A1.**
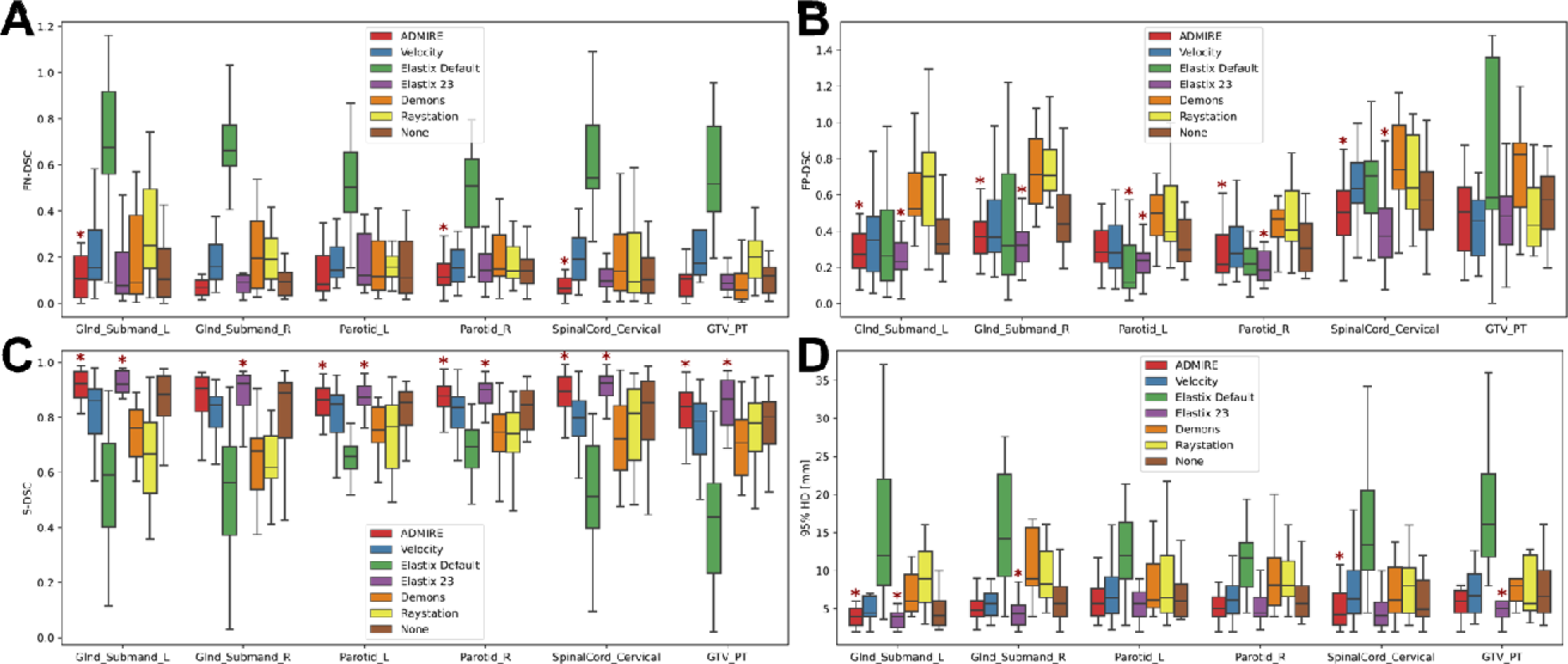
Box plots of evaluation metrics for each structure according to the registration method for false negative Dice similarity coefficient (DSC) (FN-DSC) [**A**], false negative DSC (FP-DSC) [**B**], surface DSC (S-DSC) [**C**], and 95% Hausdorff distance (95% HD) [**D**]. Asterisks indicate a significant improvement between the registration method and no registration (None). Glnd_Submand, submandibular gland; L, left; R, right; GTV_PT, primary gross tumor volume.

**Figure A2.**
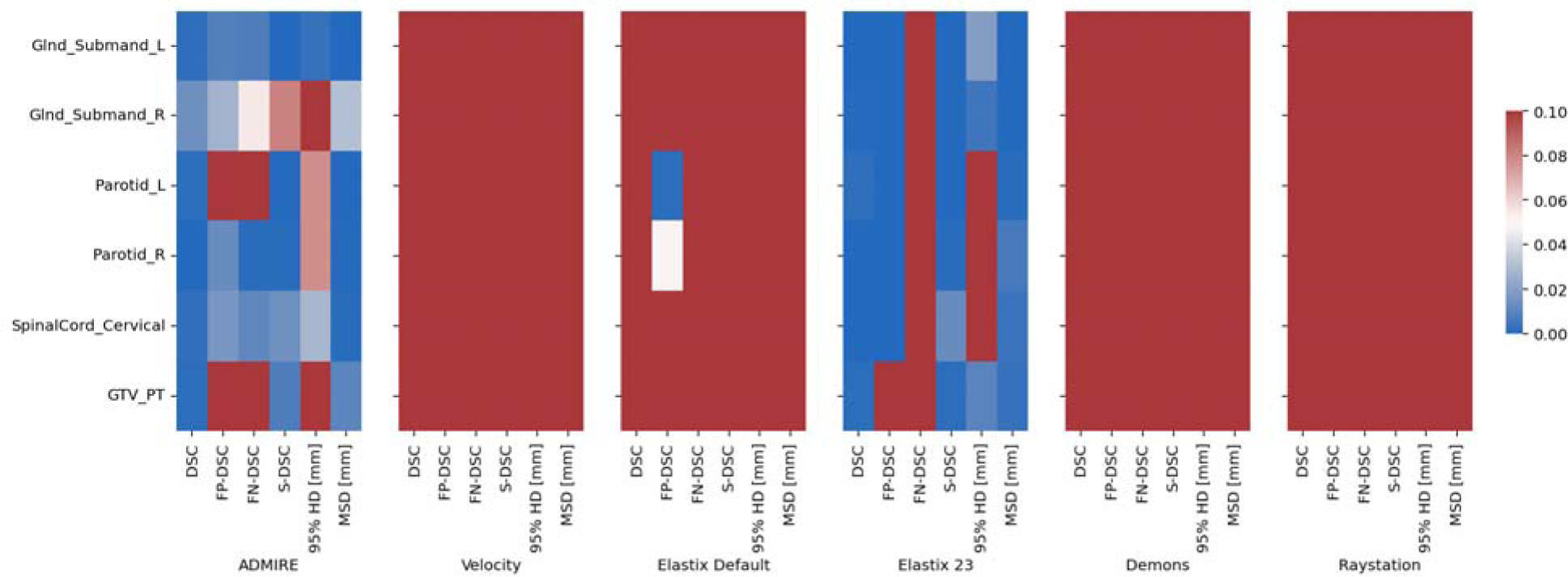
Heatmap of Bonferroni corrected p-values for one-way Wilcoxon-signed rank tests between various registration methods and no registration (None) indicating significant improvement across evaluation metrics and structures. Blue colors correspond to significant p-values (p<0.05) while red colors correspond to non-significant values (p>0.05). Glnd_Submand, submandibular gland; L, left; R, right; GTV_PT, primary gross tumor volume; DSC, Dice similarity coefficient; MSD, mean surface distance.

## Appendix B: Interobserver Variability Analysis

In a subset of five cases, segmentations for all structures in both sequences were manually generated by three additional separate observers (two physicians and one medical student) for interobserver variability analysis. Segmentations were propagated from T2W to DWI images and then compared to ground truth segmentations generated by each individual observer. For each DIR method, we implemented a Kruskal-Wallis one-way analysis of variance test ^3^ for all four observers across all structures and evaluation metrics. We performed an interobserver variability analysis to determine if there were any significant differences between observers for a given registration method. Metric value comparisons between all observers were non-significant for all structures (**Figure B1**); therefore, our study had no major interobserver variability.

**Figure B1.**
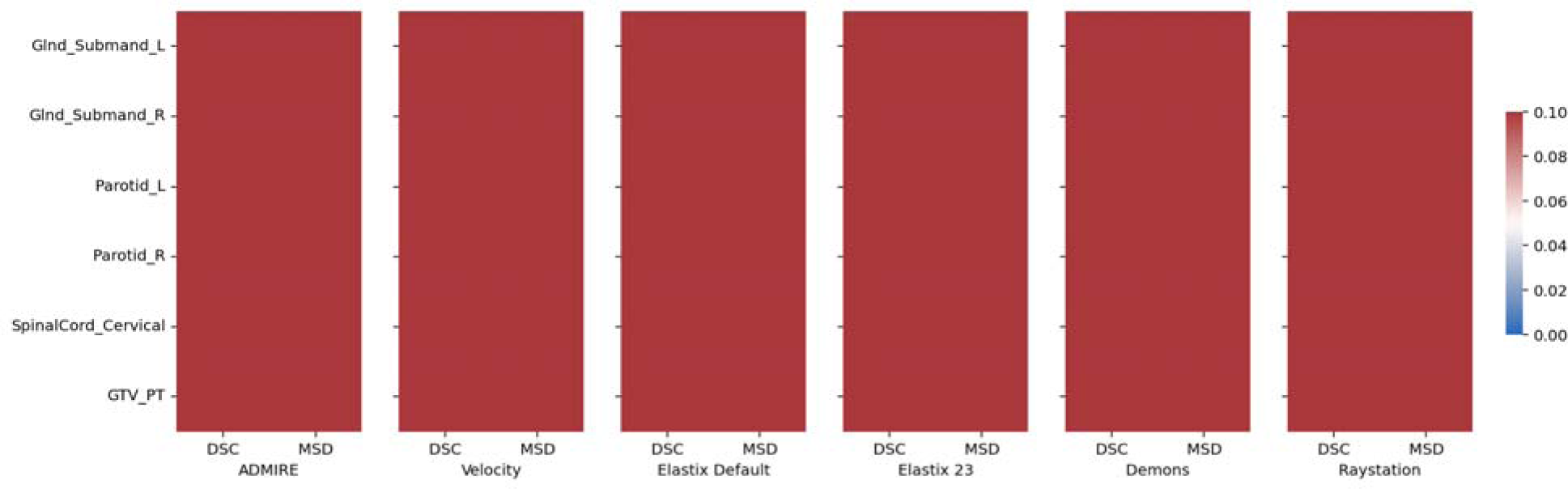
Heatmap of Bonferroni corrected p-values for one-way Wilcoxon-signed rank tests between various registration methods and no registration (None) indicating significant improvement across evaluation metrics and structures. Blue colors correspond to significant p-values (p<0.05) while red colors correspond to non-significant values (p>0.05). Glnd_Submand, submandibular gland; L, left; R, right; GTV_PT, primary gross tumor volume; DSC, Dice similarity coefficient; MSD, mean surface distance.

## Appendix C: Dosimetric Analysis

We performed a series of experiments to determine if DIR algorithms could decrease the dosimetric differences between the propagated contours and ground-truth contours. For each patient, to establish the ground-truth dose for each structure, the dose map corresponding to the radiotherapy planning computed tomography scan was propagated to the T2-weighted (T2W) image using a DIR registration in Velocity AI (v.3.0.1; Varian Medical Systems; Palo Alto, CA, USA), as has been benchmarked in previous studies ^4^. The ground-truth T2W structure contours were overlaid on the propagated dose map to determine the ground-truth dose values for each structure. For each DIR algorithm, the T2W dose map was transformed to the space of the DWI image using the corresponding deformable vector field (DVF). The previously propagated contours were then overlaid on the propagated DWI dose map to determine the propagated doses for each structure. The absolute value of the difference between the ground-truth dose and the propagated dose was then calculated for each structure (smaller values were deemed better). One-sided Wilcoxon signed rank tests (alternative hypothesis = less than) with Bonferroni corrections were then used to statistically compare DIR algorithms with no registration (None). Dose differences across all patients for the various structures and DIR algorithms are shown in **Figure C1**. A heatmap of significance values comparing DIR algorithms with None is shown in **Figure C2**.

**Figure C1.**
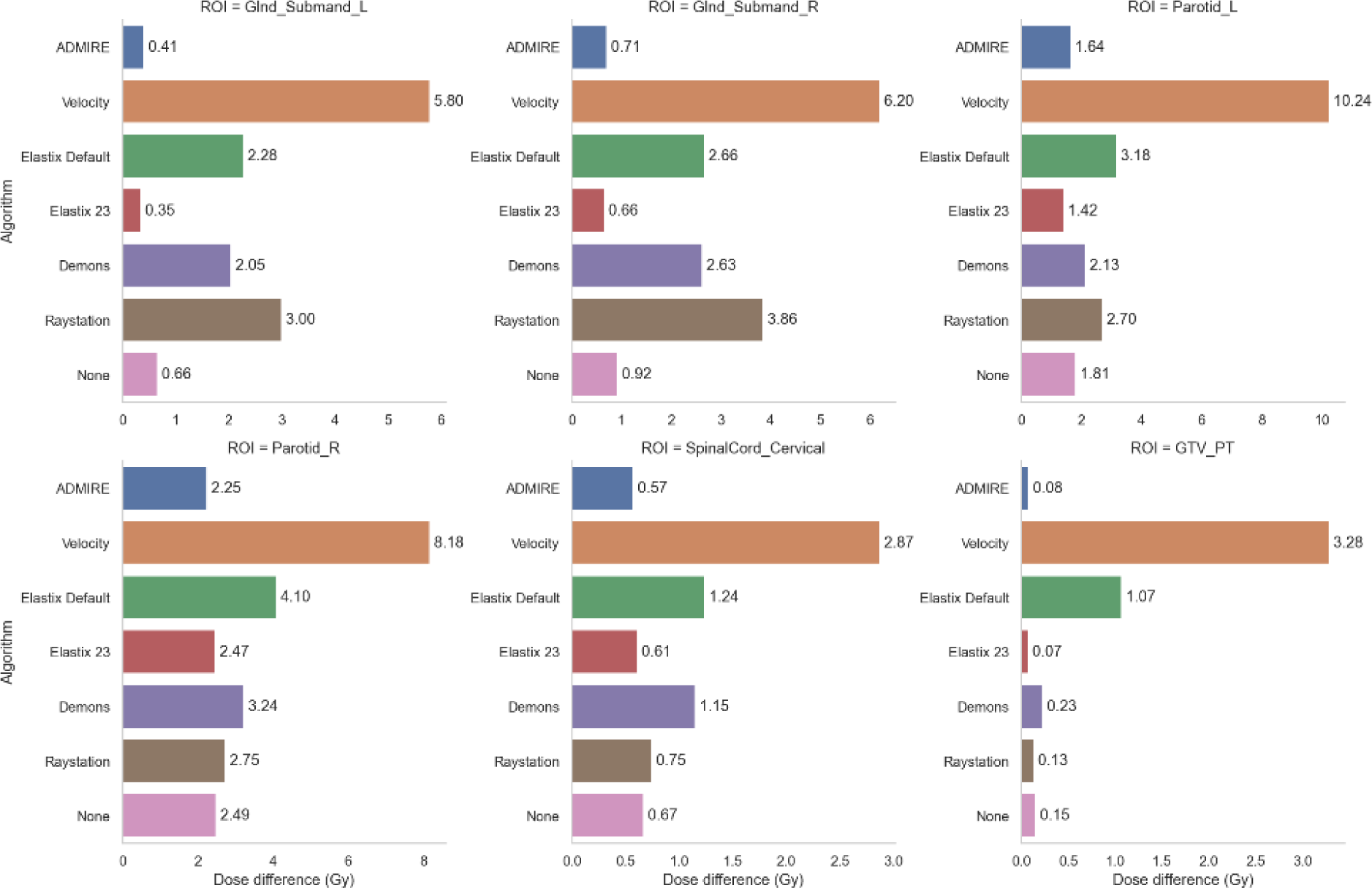
Bar plots of dosimetric differences between propagated contours and ground-truth contours based on registration method for all contoured structures. Bar value indicates mean across all patients. Glnd_Submand, submandibular gland; L, left; R, right; GTV_PT, primary gross tumor volume.

**Figure C2.**
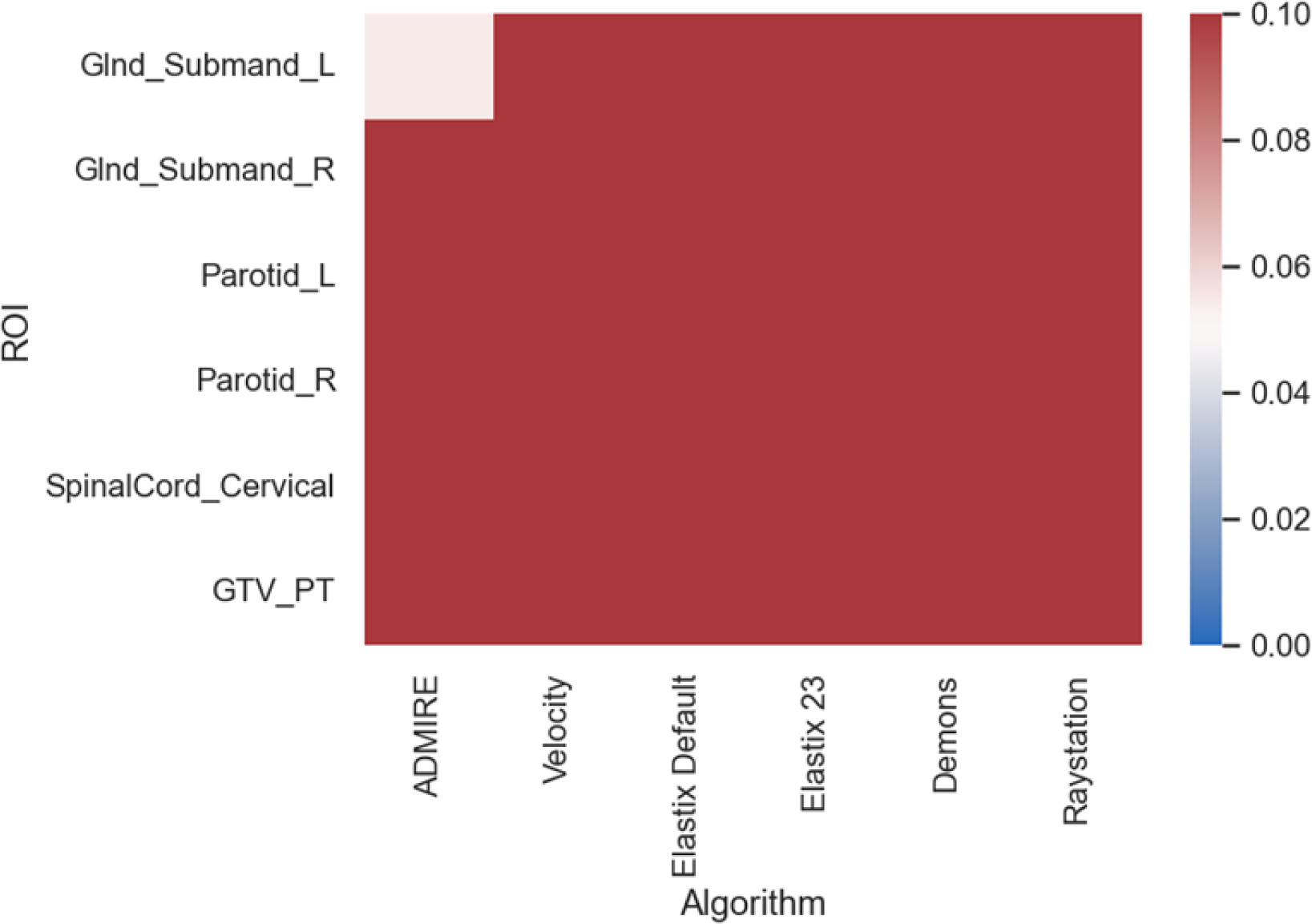
Heatmap of Bonferroni corrected p-values for one-way Wilcoxon-signed rank tests between various registration methods and no registration (None). Blue colors correspond to significant p-values (p<0.05) while red colors correspond to non-significant values (p>0.05). Glnd_Submand, submandibular gland; L, left; R, right; GTV_PT, primary gross tumor volume.

As can be seen, trends for dosimetric differences generally followed trends for geometric evaluation in the main text, i.e., ADMIRE and Elastix 23 tended to provide the best results (lowest difference values), while Velocity, Elastix Default, Demons, and Raystation tended to provide the worst results (highest difference values). However, no DIR algorithm provided statistically lower difference values when compared with None. Importantly, while the dosimetric improvements provided by ADMIRE and Elastix 23 may not be statistically significant, these dosimetric improvements may still be clinically significant. However, further studies, such as those based on normal tissue complication probability models, should be performed to confirm the clinical impact of these dosimetric differences, and are outside the scope of this study. Notably, somewhat inconsistent with the findings of the geometric evaluation, Velocity was the algorithm that led to the worst dosimetric results. However, as is reported in previous literature, dosimetric differences do not always correlate with geometric indices ^5–7^, so these results are not atypical. The excessive and at times unrealistic warping induced by the Velocity method may have led to these large dosimetric differences.

## Appendix D: Target Registration Error Analysis

To further analyze differences in image registration quality for the various DIR methods, we also performed target registration error (TRE) analysis. 6 main landmarks were identified on both T2 and DWI scans:

1. Horizontal line between the mandible angles.
2. Vertical line between the mentum and the midpoint of the anterior surface of the vertebra.
3. Bilateral medial pterygoid muscle vertical length (right).
4. Horizontal length of the cerebellum.
5. Horizontal line between the outer surface of the parotid glands.
6. Bilateral medial pterygoid muscle vertical length (left).

Landmarks were measured on the original DWI image and deformed T2W images for 3 approaches, ADMIRE, Velocity, and no registration (None) for all 20 patients. We chose to limit the number of DIR methods evaluated due to the large number of measurements needed to test all methods; ADMIRE was chosen as it was amongst the best methods geometrically, while Velocity was amongst the worst. 3D Slicer ^8^ was used to perform all measurements. The difference between the distances on the original DWI image and the deformed images were then calculated for all structures across all patients. Mann Whitney U tests were used to compare the distributions of ADMIRE and Velocity against None.

4 cases from Velocity showed excessive warping, so these cases excluded from the analysis for all cases. Certain measurements were also not included if a landmark was not visible in a given image. In total, 382 measurements were made across the various methods. The distributions of measurement differences for the various approaches are shown in **Figure D1**. Generally, median measurements ranged from less than 1 mm to 2.5 mm for most landmarks and approaches. For 5/6 landmarks (1, 2, 3, 5, 6) ADMIRE showed decreased measurement differences compared to None, but differences were not statistically significant.

**Figure D1.**
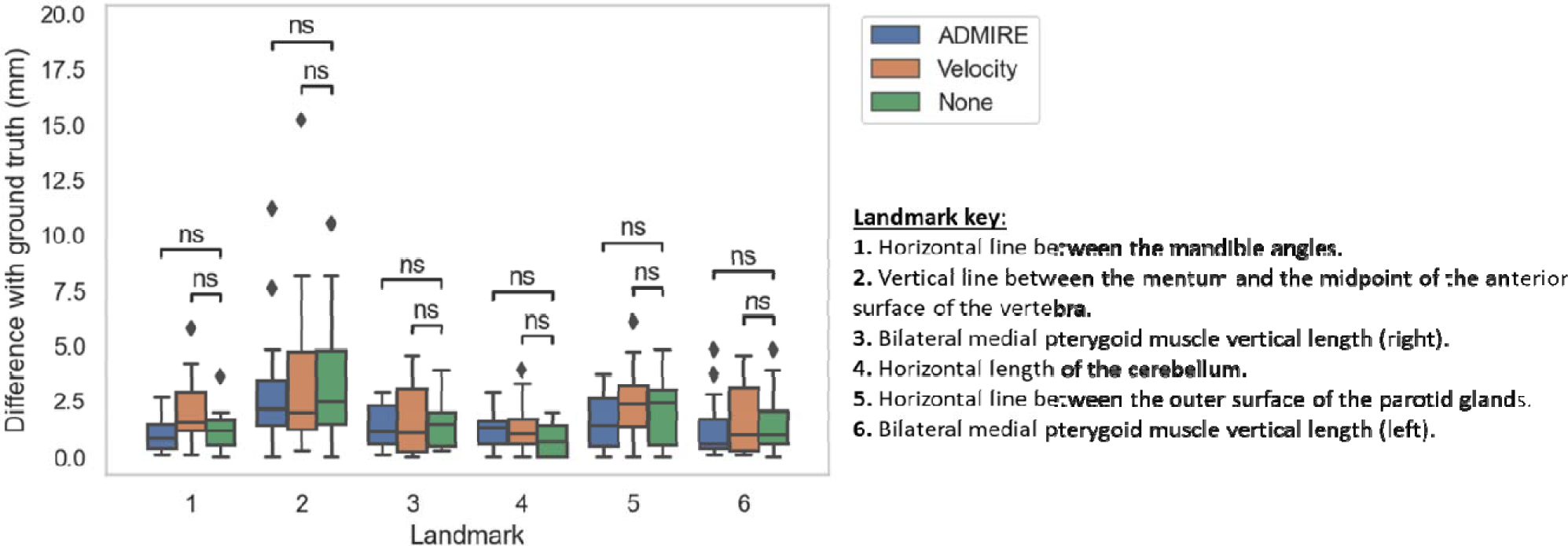
Box plots of target registration error measurements using various landmarks for 3 registration approaches (ADMIRE, Velocity, None). ns: p > 0.05 for Mann Whitney u test.

While our TRE results demonstrate some improvements for ADMIRE, unlike our geometric analysis, these results are not statistically significant. Generally, in practice it is difficult to accurately define appropriate corresponding points when registering multimodality data ^9^. Moreover, we have previously characterized TRE errors using our standard immobilization device as being less than 2 mm compared to gold-standard CT ^10^, so the small differences in deformed images may not have been easily visualized by human observers. Moreover, we believe our geometric analysis, by including surface distance measurements, is able to more robustly capture surface level details analogous to TRE measurements, and should be preferred to evaluate registration quality in this application space.

## Appendix E: Jacobian Determinant Matrix Analysis

We analyzed the Jacobian determinant matrices of the various DIR methods to evaluate the quality of the resultant DVFs. Firstly, for each method, we calculated the percentage of the negative values in the Jacobian determinant matrices (**Figure E1**). A negative Jacobian determinant indicates nonphysical motion and regions of the image folding onto itself (i.e., erroneous physical modeling of the patient) ^9^, and should therefore be avoided where possible. Velocity, Elastix Default, and Demons were the only methods with negative values for any patients, with Elastix Default having the highest amount of mean negative values (1.34%). To further investigate Jacobian determinants, we also compared the local volumetric changes based on the Jacobian integral vs. ground-truth volume differences (**Figure E2**). The Jacobian integral was defined as the mean Jacobian minus 1 times the deformed contour volume. The ground-truth volume difference was defined as the deformed contour volume minus the original T2W contour volume. Generally, the Jacobian integral measures the net local volume change, and good agreement between the Jacobian integral and volume changes indicate a reliable DVF ^11^. As shown, most algorithms showed reasonable correlation for most structures, with the exception Demons (particularly low value for primary tumor).

**Figure E1.**
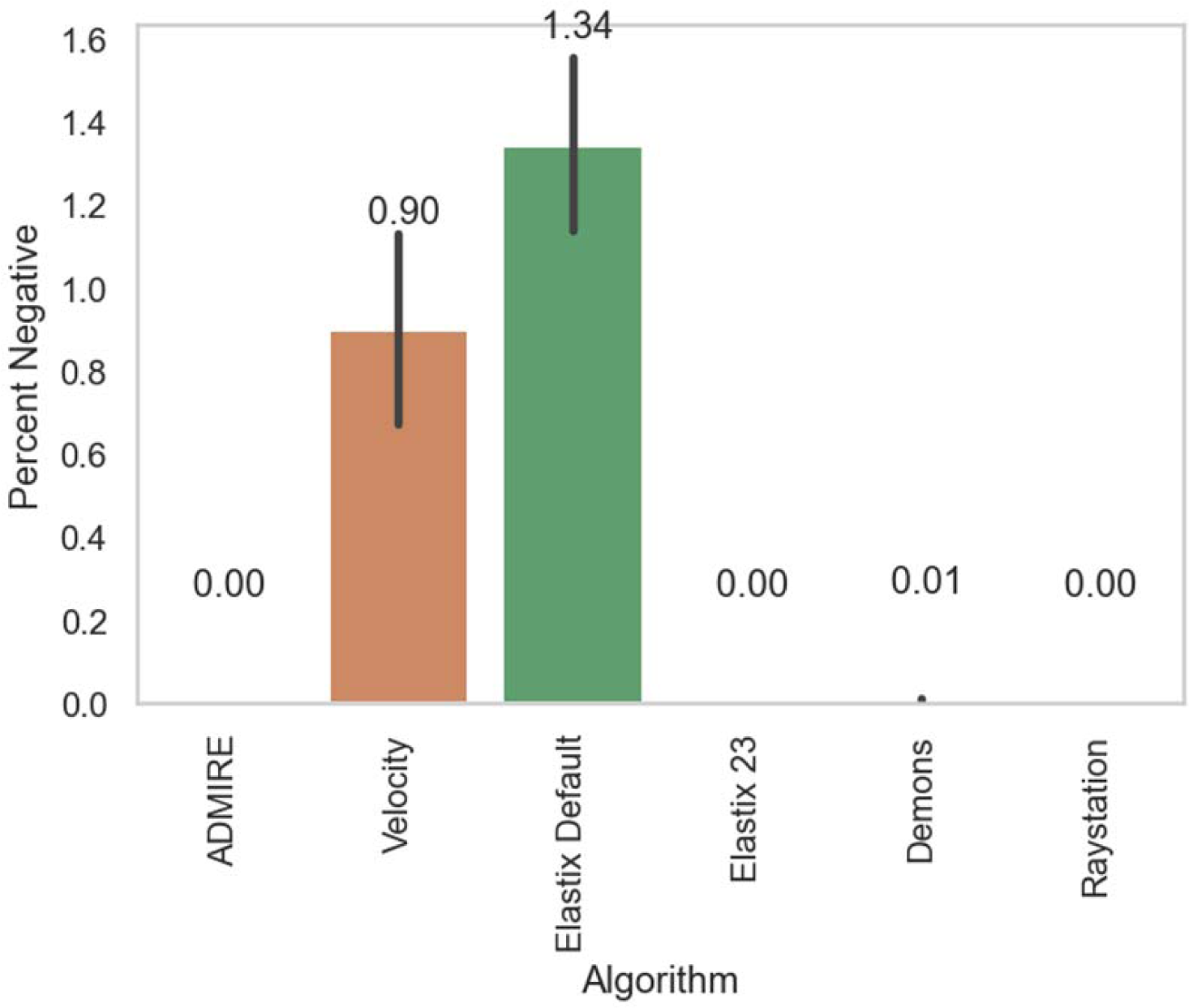
Percentage of negative values in Jacobian determinants for each deformable image registration method. Bars represent mean values across all patients (lines indicate 95% confidence interval).

**Figure E2.**
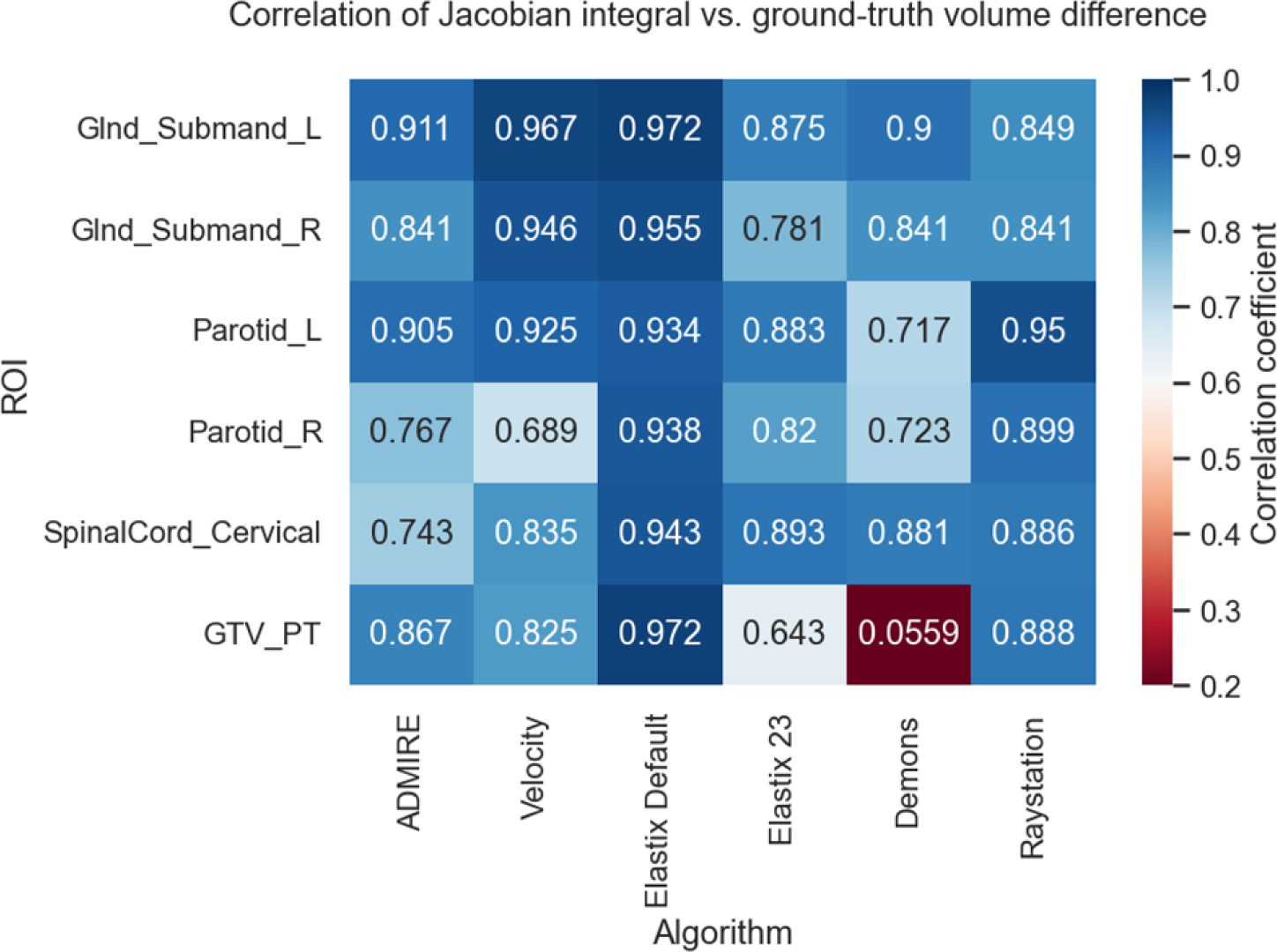
Correlation between Jacobian integral and ground-truth volume difference for various deformable image registration methods. Glnd_Submand, submandibular gland; L, left; R, right; GTV_PT, primary gross tumor volume.

## Appendix F: Spinal Cord Analysis

Of all the organ at risk (OAR) structures investigated, the cervical spinal cord had the worst overall performance across most geometric metrics, regardless of the algorithm used (**Figure 2 of main text**). Therefore, we further inspected these cases to determine if these propagated segmentations were still clinically acceptable and to determine where algorithms failed to improve overlap with the ground truth. Examples of performance of the best algorithm (ADMIRE) compared to no registration (None) and the corresponding ground truth on the diffusion-weighted image for three cases are shown in **Figure F1**. We categorized the cases as low, medium, and high, corresponding to DSC performance that was below, equal to, and above the mean performance of all cases. For the low DSC performance case, there was a clear advantage to using the ADMIRE method, as more voxels were able to overlap correctly in the superior region of the spinal cord. However, the algorithm had difficulties near the inferior portions of the spinal cord where a greater degree of curvature was present. The differences between the ADMIRE algorithm and None were less dramatic for the medium and high DSC performance cases, where there was minimal curvature in the spinal cord.

**Figure F1.**
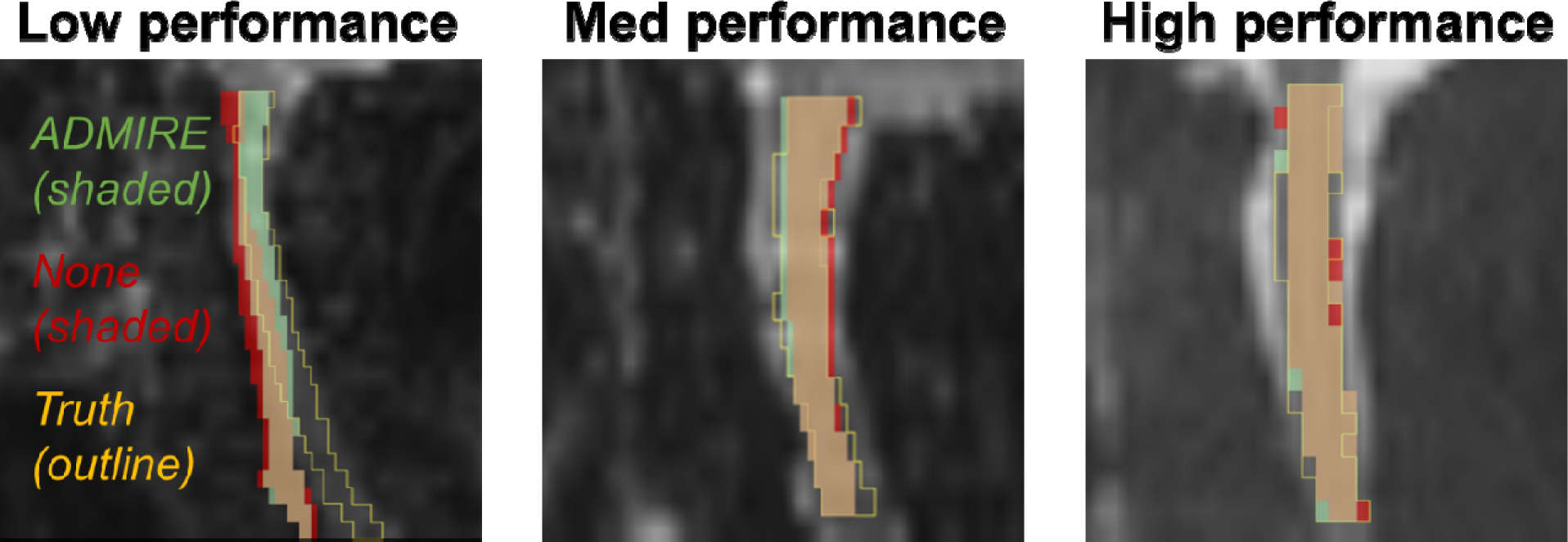
Examples of performance of the best algorithm (ADMIRE) compared to no registration (None) and the corresponding ground truth on the diffusion-weighted image for three cases (low, medium, and high DSC performance). The ADMIRE algorithm structure is shaded in green, None is shaded in red, and the ground truth structure is outlined in yellow. The overlapping areas of the ADMIRE algorithm and None are shaded in tan. DSCs of the ADMIRE algorithm and None were 0.556 and 0.335 (respectively) for the low performance case, 0.752 and 0.718 (respectively) for the medium performance case, and 0.876 and 0.815 (respectively) for the high performance case.

